# Efficacy and safety of Youth-derived Fecal Microbiota Transplantation among adults with Type 1 Diabetes Mellitus: A protocol of pilot randomized controlled trial

**DOI:** 10.64898/2026.02.03.26345459

**Authors:** Xunuo Chen, Mengyun Lei, Jun Tang, Huawei Wang, Jiawei Chen, Yanxin Liu, Shuting Li, Fang Liu, Yonge Wang, Zhiqiang Li, Zhe Dai

## Abstract

**Background:** Dysbiosis of gut microbiota plays a key role in type 1 diabetes mellitus (T1DM). Fecal microbiota transplantation represents a novel therapeutic avenue. We hypothesize that youth-derived fecal microbiota transplantation (yFMT) can remodel the gut microecosystem and improve clinical outcomes. This study aims to investigate the efficacy and safety of orally administered yFMT capsules in adults with T1DM.

**Methods and analysis:** This single-center, randomized, double-blind, placebo-controlled pilot study will enroll adults with T1DM who have suboptimal glycemic outcomes (glycated hemoglobin[HbA1c] of 7-14% and time in range [TIR] <70%). Following a 17-day run-in period for insulin optimization, continuous glucose monitoring(CGM) wearing, baseline assessments and bowel preparation, participants will be randomly allocated (1:1) to take yFMT or placebo capsules for consecutive 6 days, alongside their standard insulin therapy, and then complete a 12-week follow-up. The primary efficacy endpoint is the change from baseline in the rate of achieving the composite target of TIR>70% and time below range<4% at 4 and 12 weeks post-randomization. Secondary efficacy endpoints comprise changes from baseline at weeks 4 and 12 in other glycemic metrics (including HbA1c, fasting glucose, 2-hour postprandial glucose, and additional CGM metrics), C-peptide, immune responses, infection markers, and gut microbiota composition.

Changes from baseline at week 12 in serum metabolomic profiles will also be assessed, encompassing bile acids, short-chain fatty acids, and other related metabolites. Safety endpoints include the incidence of adverse events and serious adverse events.

**Discussion:** Our findings will offer new insight into the feasibility and effects of oral yFMT in adult with T1DM and provide the necessary evidence to power a subsequent multicenter large-scale study. Exploratory biomarker analyses conducted within this study may further pave the way for future individualized microbiome–based therapeutics.

**Trial registration:** Chinese Clinical Trial Registry identifier: ChiCTR2500111955 (November 7, 2025).

## 1. Introduction

Type 1 diabetes mellitus (T1DM) is a chronic autoimmune disease characterized by the destruction of pancreatic β cells and lifelong insulin dependence ^[1]^. Although advances in insulin analogs, delivery modalities, and continuous glucose monitoring (CGM) systems have improved diabetes management,people with T1DM still face challenges such as suboptimal glycemic outcomes, frequent episodes of hypoglycemia, and a high burden of self-management ^[2–7]^.

The core pathological mechanism of T1DM involves autoimmune-mediated destruction of pancreatic β-cell. Conventional insulin therapy fails to halt the autoimmune process or restore β-cell function. In recent years, the gut microbiota, termed the “second genome”, has been valued as a crucial role in T1DM pathogenesis ^[8]^. Numerous studies have characterized gut microbiota alterations in individuals with T1DM, marked by a reduction in butyrate-producing bacteria and secondary bile acids, along with an increase in pathobionts ^[9–11]^. Impaired intestinal barrier function represents another hallmark of this dysbiotic state, and together these features contribute to disruption of the “gut microbiota-intestinal-islet axis” ^[11]^.

Fecal microbiota transplantation (FMT) is a disruptive technique that transfers healthy donor microbiota to a recipient, aiming to restore microbial homeostasis. Evidence from animal studies suggests that modulating the gut microbiota composition can prevent or delay T1DM ^[12,13]^. Therefore, FMT holds therapeutic potential for T1DM, possibly through the following mechanisms: (1) restoration of short-chain fatty acid-producing bacteria and improvement of intestinal barrier function; (2) regulation of autoimmune responses; (3) modulation of metabolism through the gut-brain axis ^[8,14]^.

Current evidence supporting FMT in individuals with T1DM remains limited. In a randomized controlled trial, autologous but not donor-derived FMT preserved residual β-cell function for up to 12 months, with metagenomic analysis implicating changes in Parabacteroides distasonis ^[15]^. In one before-after controlled study, washed microbiota transplantation delivered via nasojejunal tube significantly improved glycemic variability in individuals with brittle diabetes ^[16]^. This improvement was associated with increased levels of lithocholic acid and isolithocholic acid, suggesting a potential mechanism involving bile acid metabolism ^[17]^. Together with two capsule-based RCTs ^[18,19]^, these studies showed that FMT was generally safe, with mild and transient gastrointestinal symptoms being the most frequent adverse events ^[15–19]^.

Despite preliminary evidence supporting the potential of FMT to treat T1DM, several challenges remain: (1)studies primarily target β-cell function with inconsistent results, and lack assessment of autoimmunity or insulin sensitivity; (2)reliance on HbA1c or isolated CGM value, while composite target of TIR(3.9-10.0 mmol/L)>70% and time below range(TBR, <3.9mmol/L)<4% are seldom used; (3)donor screening lacks unified criteria, and techniques for preparation and delivery vary widely; (4)the complex and poorly tolerated nasojejunal tube remains the primary administration method.

Therefore, our study pioneers the oral use of youth-derived FMT (yFMT) in a pilot RCT to evaluate its multidimensional effects,including glycemic outcomes, islet function, insulin sensitivity, and immune status in adults with T1DM.

## 2. Methods

### 2.1 Study Design

This is a single-center, randomized, double-blind, placebo-controlled, parallel trial conducted by Zhongnan Hospital of Wuhan University. The schedule of activities to be performed and a study flowchart are provided in **Figs 1 and 2**. Participants will be randomized in a 1:1 ratio to receive either oral yFMT capsules or an identical placebo capsule. The study will remain blinded until the database lock. Participants will provide written informed consent prior to enrollment.

**Fig 1.**
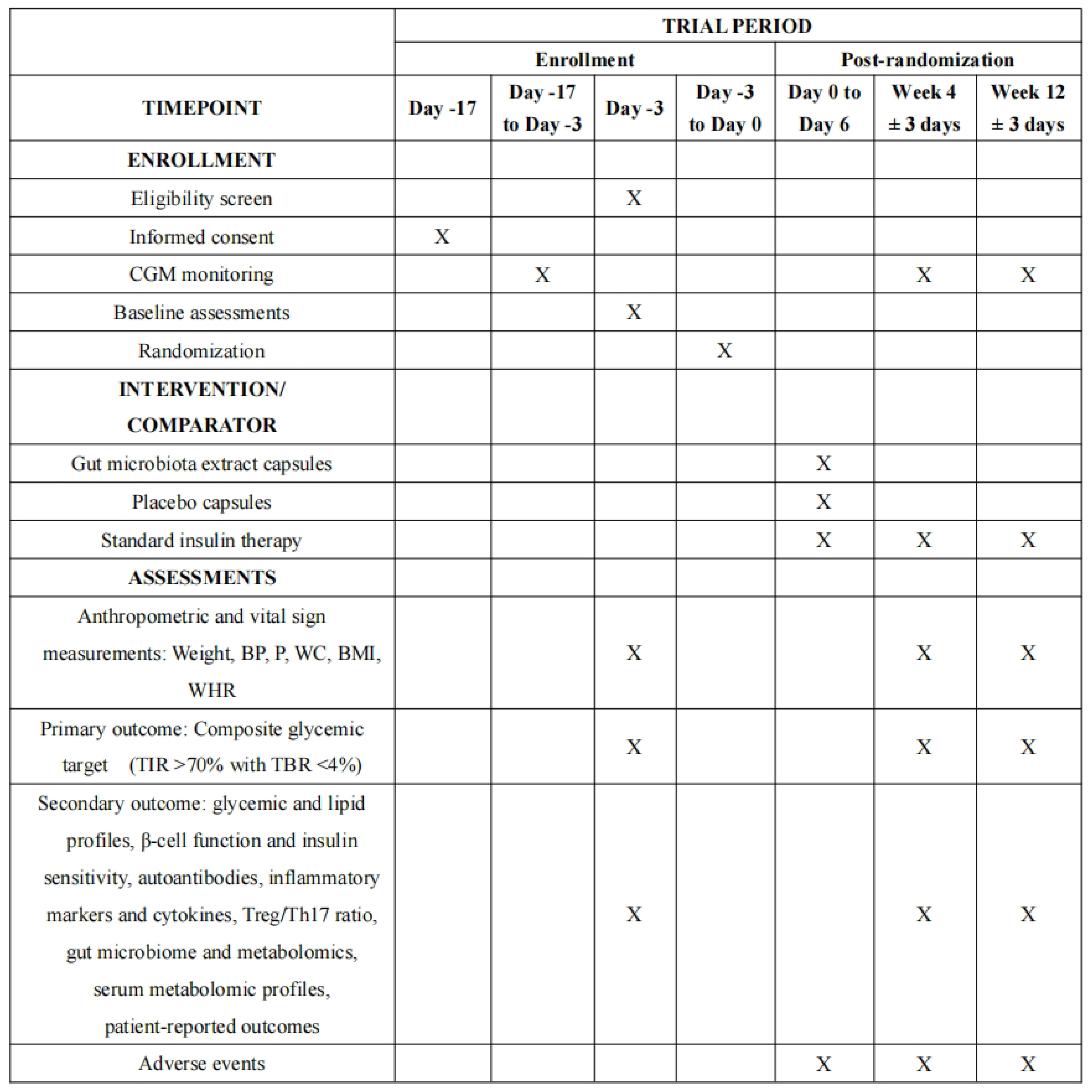
SPIRIT schedule of enrollment, intervention, and assessments. Note: Following informed consent on Day −17, candidates will undergo a 14-day CGM assessment and HbA1c testing (Day −3). Only those with HbA1c of 7.0-14.0% or CGM-derived TIR below 70% will be enrolled. Abbreviations: CGM, Continuous Glucose Monitoring; BP, Blood Pressure; P, Pulse; WC, Waist Circumference; BMI, Body Mass Index; WHR, Waist-to-Hip Ratio; TIR, Time in Range; TBR, Time Below Range; Treg, Regulatory T cells; Th17, T helper 17 cells;

**Fig 2.**
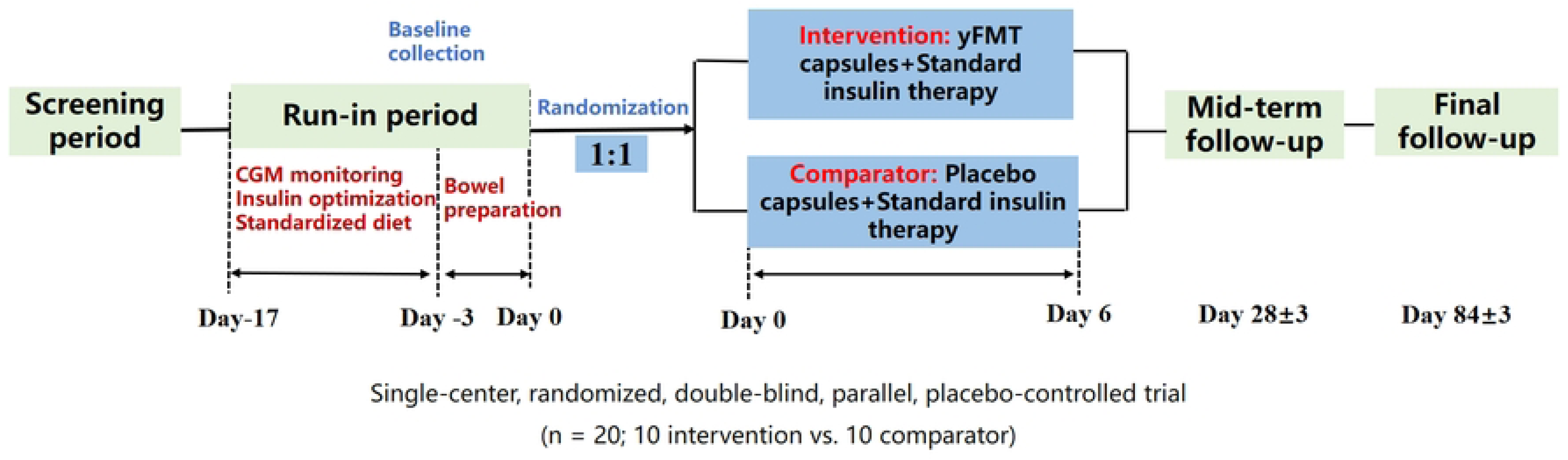
Flow chart of design. Abbreviations: CGM, Continuous Glucose Monitoring; yFMT, youth-derived fecal microbiota transplantation;

### 2.2 Trial status

This study was approved by the Medical Ethics Committee, Zhongnan Hospital of Wuhan University (Ethics Approval No. 2025302) in October 2025. Recruitment started on November 20, 2025, and is ongoing, with completion expected by July 31, 2026. Data collection will conclude by November 30, 2026, with results expected by January 31, 2027.

### 2.3 Participants

Participant recruitment will be conducted from November 2025 to July 2026. Eligible participants aged 18-60 years must have over one year duration of T1DM and suboptimal glycemic outcomes (HbA1c of 7.0-14.0% or TIR<70%). Participants will be excluded if they had recent episodes of diabetic ketoacidosis, concurrent severe gastrointestinal diseases, significant gastrointestinal diseases or surgery, pregnancy, or lactation. The diagnosis of T1DM adheres to the criteria established by the World Health Organization and the American Diabetes Association. Full criteria are listed in **Table 1**.

**Table 1.** Inclusion and exclusion criteria for enrolment.

| Inclusion Criteria |  |
| --- | --- |
| 1 | T1DM was diagnosed by an endocrinologist for at least 1 year |
| 2 | Aged from 18 to 60 years |
| 3 | HbA1c 7.0-14% or TIR<70% |
| 4 | On MDI or insulin pump therapy for $\geq 3$ months |
| 5 | TDD before admission $\geq 0.3$ IU/kg/day, with a basal rate $\geq 0.05$ IU/hour |
| 6 | Women of childbearing age are willing to use appropriate contraceptive measures |
| 7 | Signed informed consent form |
| Exclusion Criteria |  |
| 1 | Treatment with any anti-hyperglycemic medications other than insulin |
| 2 | Continuous use of antimicrobial agents, probiotics, intestinal microecological modulators, or other medications with potential impact on gut microbiota for >3 days within 3 months prior to enrollment; |
| 3 | Occurrence of acute diabetic complications (diabetic ketoacidosis, hyperosmolar hyperglycemic state) within 1 month prior to enrollment; |
| 4 | Coexisting gastrointestinal diseases (including persistent vomiting, confirmed/suspected gastrointestinal obstruction, short bowel syndrome, inflammatory bowel disease, active major gastrointestinal bleeding, gastrointestinal perforation, high-output intestinal fistula, severe diarrhea, significant fibrotic intestinal stenosis, fulminant colitis, or toxic megacolon), history of gastrointestinal surgery (such as gastrectomy, pelvic anastomosis, colostomy, or other digestive system procedures), malignancy, or sepsis; |
| 5 | Coexisting severe cardiovascular or cerebrovascular diseases, hepatic or renal disorders, hematopoietic system diseases, severe immunosuppression, or congenital/acquired immunodeficiency diseases; |
| 6 | Recent administration of high-risk immunosuppressive or cytotoxic medications: e.g., rituximab, doxorubicin, or medium-to-high dose corticosteroids (prednisone $\geq 20$ mg/day) for more than 4 weeks; |
| 7 | Use of closed-loop systems or sensor-augmented pumps within the past 3 months; |
| 8 | Concurrent participation in another randomized clinical trial; |
| 9 | Pregnancy, lactation, or planned pregnancy during the study period in female participants; |
| 10 | Inability to adhere to the recommended intervention and follow-up schedule for any reason. |

**Abbreviations:** T1DM, type 1 diabetes mellitus; HbA1c, glycated hemoglobin; MDI, multiple daily insulin injections; TDD, total daily insulin doses.

### 2.4 Procedures

#### 2.4.1 Screening (Day −17)

Screening (Day −17, Outpatient): Informed consent will be obtained, and eligibility will be assessed against all inclusion and exclusion criteria. Eligible participants will be provided with a CGM device.

#### 2.4.2 Run-in Period and Baseline Assessments (Day −17 to Day 0)

This period standardizes participants’ status prior to intervention and consists of two phases:

*Outpatient Phase* (Day −17 to −3): Participants will wear CGM, undergo insulin dose optimization and receive dietary guidance according to the Chinese Diabetes Diet Guidelines (2017). CGM data from this phase serves as the baseline for glycemic metrics, including TIR, TBR, glucose management indicator (GMI), coefficient of variation (CV), and other metrics.

*Inpatient Phase* (Day −3 to 0): Participants are hospitalized for baseline assessments and bowel preparation:

1. Baseline Assessments (Completed by Day −3): including:

a. Demographics and clinical parameters: age, sex, T1DM duration, anthropometrics (height, weight, waist-to-hip ratio [WHR]), vital signs (blood pressure, pulse), and comprehensive medical/medication history;
b. Glycemic and lipid profiles: fasting plasma glucose, 2-hour postprandial glucose via mixed meal test[MMT], HbA1c, total cholesterol (TC), triglycerides (TG), low-density lipoprotein cholesterol [LDL-c], and high-density lipoprotein cholesterol [HDL-c]);
c. β-cell function and insulin sensitivity (fasting/stimulated 2-hour C-peptide during Mixed-Meal Tolerance Test [MMT 2h CP], estimated glucose disposal rate[eGDR]);
d. Diabetic autoantibodies: Glutamic Acid Decarboxylase Autoantibodies [GADA], Insulinoma-Associated protein 2 Autoantibodies [IA-2A], Zinc Transporter 8 Autoantibodies [ZnT8A]);
e. Inflammatory markers: white blood cell count [WBC], C-reactive protein [CRP], erythrocyte sedimentation rate [ESR], procalcitonin [PCT], and cytokines [TNF-α, IL-1β, IL-6, IL-8, IFN-γ]);
f. Immune marker:Treg/Th17 ratio
g. Gut microbiome and serum metabolomics;
h. Patient-reported outcomes (PROs): Simplified Chinese version of the EQ-5D-5L, Diabetes Distress Scale [DDS], Chinese version of the Hypoglycemia Fear Survey II-Worry Scale [CHFSII-WS]);
i. Laboratory safety indicators: liver and renal function.
2. Bowel Preparation: from Day −3, participants receive oral rifaximin (0.2 g per dose, four times daily), with bowel preparation using a polyethylene glycol solution performed on Day 0.

#### 2.4.3 Randomization and Blinding

Following the completion of the run-in period, eligible participants will be randomized in a 1:1 ratio to either the intervention or the control group. Randomized allocation sequences will be generated using a permuted block design in R software. Sequentially numbered, opaque, sealed envelopes will be prepared and stored by a non-investigational team member. To minimize selection bias, the randomization codes are assigned by an independent researcher. Once an eligible participant is enrolled, the principal investigator notifies the independent researcher to open sequentially the sealed envelope for allocation. yFMT and placebo capsules are identical in appearance, shape, odor, and specifications.

The investigational capsules will be assigned predefined random codes that were dispensed without selection. Randomization number is not allowed to be reassigned following participant discontinuation. Blinding will be maintained for both participants and endpoint assessors throughout the study. The blinding code will be held by an independent statistician securely until the final analysis.

#### 2.4.4 Study intervention

1. Regimen: yFMT capsules is derived from healthy donors aged 4-17 years, who capture the peak period of gut microbiota diversity. A three-dimensional matching system based on disease status, sex, and age was established to achieve precise donor-recipient matching for the preparation of oral gut microbiota capsules (**Fig 3**). Eligible participants will be randomized to receive either yFMT capsules (containing≥ 4×10¹³ live bacteria) or identical capsules (skimmed milk and starch). The administration protocol is as follows: 6 capsules are to be taken orally twice daily (a total of 12 capsules/day) under fasting conditions (at least 2 hours before or after meals) for six consecutive days. All capsules must be swallowed whole without chewing to maintain bacterial viability and blinding. All participants maintain their usual insulin therapy throughout the study period and are allowed to adjust their insulin dose based on CGM data.
2. Storage and Handling: Capsules are stored at −80°C and must be administered within 20 minutes of thawing.
3. Safety Monitoring: During the 6-day intervention period, participants will be monitored daily for:

a. Gastrointestinal tolerance: specifically assessing bloating, abdominal pain, changes in bowel habits (constipation or diarrhea), nausea, or vomiting.
b. Systemic symptoms: fever, rash, or any new allergic-like reactions.
c. Glucose metabolism: close review of CGM data for significant or unexplained glycemic fluctuations, particularly hypoglycemia.

Any adverse events (AEs) will be recorded, and their relationship to the study intervention will be assessed using a predefined causality algorithm specific to microbiota-based therapies.

**Fig 3.**
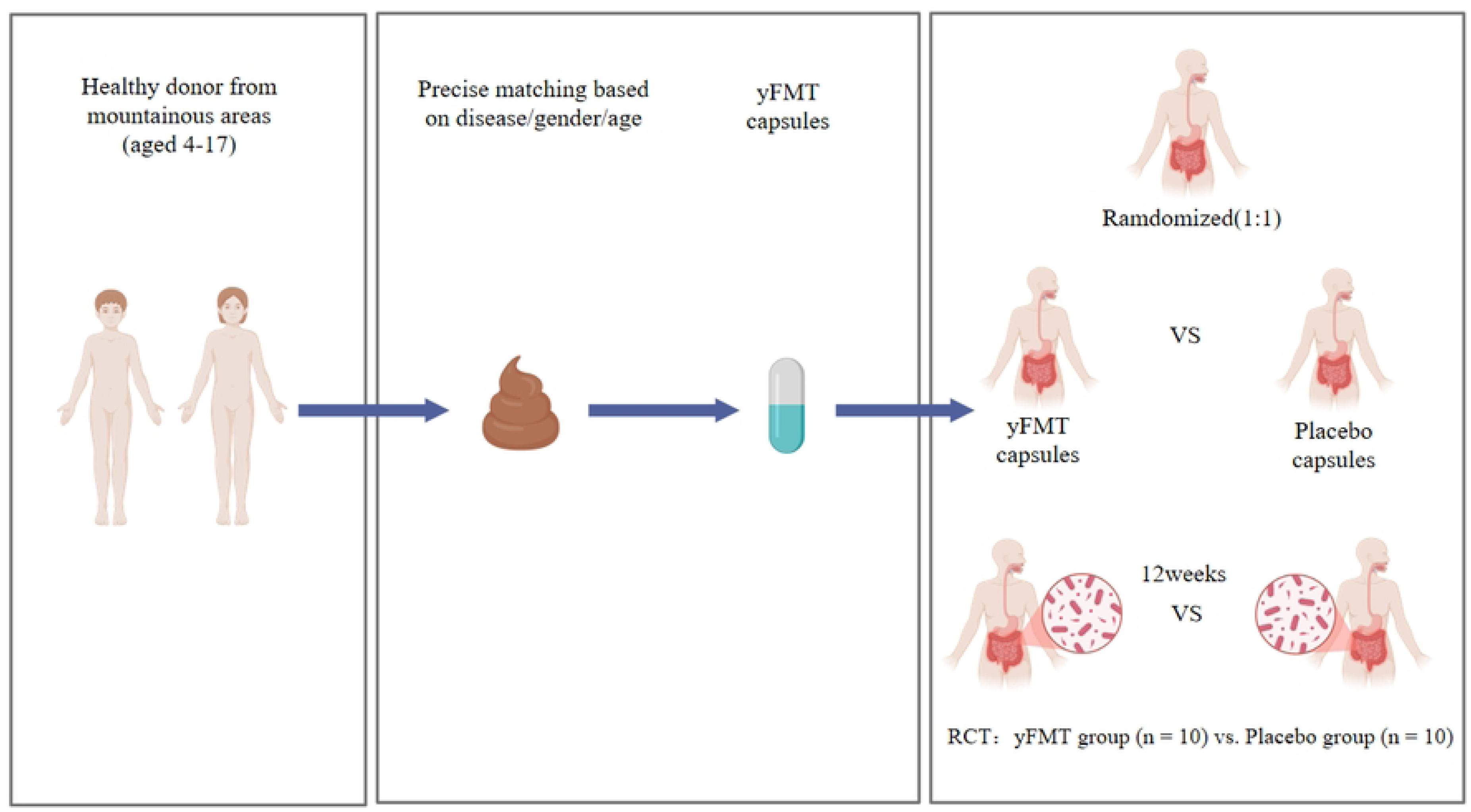
Source and allocation of gut microbiota capsules. Abbreviations: yFMT, youth-derived fecal microbiota transplantation; RCT, Randomized Controlled Trial;

#### 2.4.5 Follow-Up Visits

1. Interim Visit (Week 4 ±3 days, outpatient): Includes clinical evaluation, laboratory tests (metabolic, β-cell function, immune markers), CGM data collection (Weeks 2-4), PROs, and AEs/severe adverse events (SAE) documentation.
2. Final Visit (Week 12 ±3 days, outpatient): A comprehensive reassessment, including all parameters from the interim visit plus final gut microbiome and safety laboratory samples. CGM data from Weeks 10-12 are collected. The study concludes with database lock and unblinding.

A detailed schedule is provided in **Table 2**.

**Table 2.** Study Visit Schedule.

| Process | Screening period | Run-in period 1 | Baseline period | Run-in period 2 | Intervention period | Mid-term follow-up | Final follow-up |
| --- | --- | --- | --- | --- | --- | --- | --- |
| | Day -17 | Day -17 to Day -3 | Day -3 | Day -3 to Day 0 | Day 0 to Day 6 | Week 4 ± 3 days | Week 12 ± 3 days |
| Obtain informed consent | √ |  |  |  |  |  |  |
| Basic characteristics | √ |  |  |  |  |  |  |
| Lifestyle survey |  |  | √ |  |  |  |  |
| Record anthropometric measurements |  |  | √ |  |  | √ | √ |
| Average total daily insulin dose (Recent 1-2 Weeks) |  |  | √ |  |  | √ | √ |
| Glucose + C-peptide ( Fasting and MMT 2h ) |  |  | √ |  |  | √ | √ |
| HbA1c | √ |  | √ |  |  | √ | √ |
| Islet autoantibody titer |  |  | √ |  |  | √ | √ |
| Treg/Th17 ratio |  |  | √ |  |  | √ | √ |
| Lipids profile |  |  | √ |  |  | √ | √ |
| Liver function |  |  | √ |  |  |  | √ |
| Renal function |  |  | √ |  |  |  | √ |
| Thyroid function (five items) |  |  | √ |  |  |  | √ |
| Infection markers and inflammatory cytokines |  |  | √ |  |  | √ | √ |
| Safety assessments |  |  | √ |  | √ | √ | √ |
| AEs and SAEs |  |  | √ |  |  | √ | √ |
| Gut microbiome |  |  | √ |  |  | √ | √ |
| Serum metabolomics |  |  | √ |  |  |  | √ |
| Oral + bowel cleansing |  |  |  | √ |  |  |  |
| Oral yFMT capsules |  |  |  |  | √ |  |  |
| Standard insulin therapy |  | √ | √ | √ | √ | √ | √ |
| CGM monitoring |  | √ |  |  | √ | √ | √ |
| Questionnaires |  |  | √ |  |  | √ | √ |
| Provision of yFMT to the placebo group after unblinding |  |  |  |  |  |  | √ |

**Note:** Basic characteristics: age, sex, ethnicity, height, weight, waist circumference, hip circumference, duration of T1DM, past medical history, and medication history; Lifestyle survey: smoking, alcohol consumption, diet, and physical activity; Record anthropometric measurements: height, weight, blood pressure, pulse, waist circumference, hip circumference, BMI, and WHR; Infection markers: WBC, ESR, CRP, PCT; Inflammatory cytokines: TNF-α , IL-1β , IL-6, IL-8, IFN-γ ; Safety assessments: body temperature, pulse, respiration, and blood pressure; Adverse events and serious adverse events: including abnormalities in liver and kidney function tests, as well as symptoms such as nausea, abdominal pain, diarrhea, abdominal distension, fatigue, constipation, loss of appetite, and allergic reactions. SAEs refer to medical events occurring during the clinical trial that result in hospitalization or prolonged hospitalization, disability, impaired work capacity, life-threatening conditions, death, or congenital malformations; Completion of relevant questionnaires: Simplified Chinese version (Mainland China) of the EQ-5D-5L scale, Chinese version of the Hypoglycemia Fear Survey-Worry Subscale (HFS-WS), and the Diabetes Distress Scale (DDS).
**Abbreviations:** MMT 2h, Mixed-Meal Tolerance Test, 2 hours; HbA1c, glycated hemoglobin; Treg, Regulatory T cell; Th17, T helper 17 cells; yFMT, youth-derived Fecal Microbiota Transplantation; CGM, Continuous Glucose Monitoring; T1DM, Type 1 Diabetes Mellitus; BMI, Body Mass Index; WHR, Waist-to-Hip Ratio; WBC, White Blood Cell; ESR, Erythrocyte Sedimentation Rate; CRP, C-reactive Protein; PCT, Procalcitonin; TNF-α , Tumor Necrosis Factor-alpha; IL-1β , Interleukin-1 $\beta$ ; IL-6, Interleukin-6; IL-8, Interleukin-8; IFN-γ , Interferon gamma.

#### 2.4.6 Post-Study Procedures

Participants from the placebo group will be offered an optional course of open-label yFMT treatment. All participants will retain access to a 24-hour safety hotline for reporting any subsequent AEs.

### 2.5 Endpoints

The primary endpoint was the composite achievement rate of TIR>70%+TBR<4% . Secondary efficacy endpoints included glycemic metabolism, β-cell function, insulin sensitivity, immunomodulation, inflammatory markers, gut microbiome and metabolites, PROs, lipid profile and TDD. Safety endpoints comprised the incidence of AEs (such as gastrointestinal symptoms, infection-related events, immune-related reactions, and allergic responses), as well as the incidence of serious AEs (SAEs, including severe hypoglycemia, diabetic ketoacidosis, hospitalization, disability, life-threatening events, or death). Detailed information is provided in **Table 3**.

**Table 3.** Study Endpoints.

|  |  |
| --- | --- |
| **Primary endpoint** | Changes in the composite glycemic target achievement rate (TIR >70% with TBR <4%) |
| **Secondary endpoints** | Changes in glycemic parameters ( including HbA1c, FBG, MMT 2h PG and other CGM metrics [TIR, TBR, TAR, GMI, GRI, CV]). |
| | Changes in pancreatic β-cell function (including FCP, MMT 2h CP, and the AUC of MMT 2h CP) |
|  | Changes in insulin sensitivity indicators (including eGDR) |
|  | Changes in TDD |
|  | Changes in infection and inflammatory markers (including WBC, PCT, ESR, CRP, and inflammatory cytokines) |
|  | Changes in cellular immune indicators (including peripheral blood Treg/Th17 ratio and islet-related autoantibodies [GADA, IA-2A, ZnT8A]) |
|  | Changes in lipid profile (including TC, TG, LDL-c, and HDL-c) |
|  | Changes in gut microbiota composition and abundance |
|  | Changes in serum metabolomic profiles (including bile acids [e.g., UDCA, TUDCA] and SCFAs) |
|  | Changes in PROs ( including EQ-5D-5L, CHFSII-WS, and DDS ) |
| **Safety endpoints** | Number of serious adverse events (including severe hypoglycaemia, diabetic ketoacidosis and severe allergic reactions) |

**Note:** All changes in the above-mentioned indicators are expressed relative to baseline values. Changes in serum metabolomic profiles is assessed only at baseline and at week 12. All other indicators are evaluated at baseline, week 4, and week 12.
**Abbreviations:** TIR, Time in Range; TBR, Time Below Range; HbA1c, Glycated haemoglobin; FBG, Fasting Blood Glucose; MMT 2h PG, 2-hour Postprandial Glucose during Mixed-Meal Tolerance Test; CGM, Continuous Glucose Monitoring; TAR, Time Above Range; GMI, Glucose Management Indicator; GRI, Glycemic Risk Index; CV, Coefficient of Variation; FCP, Fasting C-Peptide; MMT 2h CP, 2-hour Postprandial C-Peptide during Mixed-Meal Tolerance Test; AUC, Area Under the Curve; eGDR, Estimated Glucose Disposal Rate; TDD, Total daily insulin doses; WBC, White Blood Cell count; PCT, Procalcitonin; ESR, Erythrocyte Sedimentation Rate; CRP, C-Reactive Protein; Treg, Regulatory T cells; Th17, T helper 17 cells; GADA, Glutamic Acid Decarboxylase Antibody; IA-2A, Insulinoma-Associated Antigen-2 Antibody; ZnT8A, Zinc Transporter 8 Antibody; TC, Total Cholesterol; TG, Triglycerides; LDL-c, Low-Density Lipoprotein Cholesterol; HDL-c, High-Density Lipoprotein Cholesterol; UDCA, Ursodeoxycholic Acid; TUDCA, Tauroursodeoxycholic Acid; SCFAs, Short-Chain Fatty Acids; PROs, patient-reported outcomes; EQ-5D-5L, EuroQol Five-Dimension Five-Level Questionnaire; CHFSII-WS, Chinese version of the Hypoglycemia Fear Survey II–Worry Scale; DDS, Diabetes Distress Scale;

### 2.6 Risk and AEs

An AE is defined as any unfavorable medical occurrence in an individual after signing the informed consent form and entering the study until the last follow-up, regardless of whether it had a causal relationship with yFMT capsules. The anticipated risks associated with included gastrointestinal symptoms (e.g., nausea, abdominal pain, diarrhea, bloating, constipation), fatigue, decreased appetite, and potential metabolic or immune-related complications (e.g., diabetic ketoacidosis, severe hypoglycemia, severe allergic reactions).

In the event of any AEs, the investigators will provide prompt management and may discontinue the study intervention based on the clinical judgment of the investigator. All AEs, regardless of severity, will be recorded in the case report form and followed until satisfactory resolution. Only SAEs are required to be reported to the Institutional Review Board as mandated by the protocol and regulatory guidelines.

### 2.7 Withdrawal standard

A data safety monitoring board will periodically convene to make clinical judgments on who should withdraw from the study. The following withdrawal rules will be abided by (1) SAEs, (2) failure to complete the yFMT intervention per protocol (e.g., missing ≥50% of the doses), (3) pregnancy or planned pregnancy, and (4) development of a newly diagnosed condition that met exclusion criteria (e.g., inflammatory bowel disease, malignancy) or any other situation in which the investigator determined that study discontinuation was warranted. It should be noted that participants maintain the right to terminate the study at any time without impact on future care.

### 2.8 Data management

All pseudonymized research data, including demographics, laboratory data, questionnaire responses, and AEs records, will be recorded in the case report form and securely maintained at Zhongnan Hospital of Wuhan University under strict confidentiality and data security protocols.

To ensure data integrity and accessibility, the following measures will be implemented: (1) all investigators will receive standardized training on data management procedures prior to trial initiation; (2) responsible investigators will perform ongoing monitoring and periodic quality checks to assess data integrity throughout the study; (3) the principal investigator will oversee the data lifecycle and audit procedures.

### 2.9 Sample Size and Pilot Trial Rationale

This investigation is a pilot study. Its primary aim is not to establish the efficacy of yFMT, but to address the key uncertainties in this novel interventional field to inform the design of a future definitive clinical trial. The specific pilot objectives are to assess the feasibility (recruitment, adherence) and safety of the oral yFMT protocol, to evaluate the variability of the primary outcome (composite glycemic target achievement rate), and to collect preliminary effect-size estimates for the planning of a subsequent pivotal trial.

As no prior data exist on achieving the composite glycemic target (TIR >70% with TBR <4%) following yFMT in adults with T1DM, a traditional sample size calculation based on an anticipated effect size was not feasible. The sample size for this trial was therefore determined in accordance with methodological best practices for pilot trial design. We referenced the guidance proposed by Wang et al., which indicates that a pilot study for a future definitive trial with 90% power and a two-sided alpha of 0.05 typically requires approximately 10 to 25 participants per group to provide meaningful parameter estimates for small-to-moderate effect sizes ^[20]^. Based on this, and considering the common requirement for assessing primary feasibility objectives (e.g., recruitment rate, procedural acceptance), a sample size of 10 participants per group was set.

The pre-planned interim analysis in this protocol will allow for a careful assessment of the accumulating feasibility and safety data to determine whether an adjustment to the final sample size is warranted.

### 2.10 Statistics and Analysis

Efficacy analyses will be performed in the intention-to-treat population, with participants missing the final assessment classified as non-responders. Safety analyses will be performed in the safety analysis set. For continuous variables, normality will be assessed using the Shapiro-Wilk test. Data will be summarized as mean ± standard deviation, or median (Q1, Q3) and analyzed using independent samples t-test/Mann-Whitney U test (between-group) or the paired t-test/Wilcoxon signed-rank test (within-group), as appropriate. Categorical data will be expressed as count (percentages) and analyzed using Fisher’s exact test.

The primary efficacy endpoint (the achievement rate of composite glycemic target) and categorical secondary endpoints will be analyzed using generalized estimating equations to assess the effects of group, time, and interaction effects. Continuous secondary endpoints will be analyzed using a linear mixed-effects model, including fixed effects for group, time, interaction, and baseline value, and a random intercept for subject. Missing data for the primary endpoint will be handled using the last observation carried forward method. Logistic regression analysis will be used to explore factors associated with response to yFMT treatment. CGM metrics, as recommended by international expert consensus, will be calculated using Glyculator 2.0 software (Medical University of Lodz, Poland). All statistical analyses will be performed using R software (version 4.3.2) and GraphPad Prism 9.0. Statistical significance is defined as two-sided P<0.05. Bonferroni correction will be applied for multiple comparisons in post hoc analyses.

## Discussion

T1DM remains a major clinical challenge as standard insulin therapy fails to halt the underlying autoimmune destruction of pancreatic β cells. The well-established association between T1DM and gut dysbiosis, supported by preclinical studies in which microbial modification slowed disease progression, has positioned FMT as a viable therapeutic strategy ^[9]^. Recent pioneering human trials have further indicated that FMT may help preserve residual islet function and improve glycemic outcomes ^[16,17]^, yet several methodological constraints impede its translation.

Current limitations include the reliance on invasive, poorly–tolerated nasojejunal tubes for delivery-which is a key barrier to clinical adoption-and the absence of standardized donor screening, which yields inconsistent microbial compositions. Furthermore, outcome are typically assessed using isolated glycemic metric such as HbA1c, fasting blood glucose, or isolated CGM metrics rather than integrated metabolic profiling.

To address these gaps, we propose a pilot study of yFMT capsules. This study features transplants from a meticulously selected pediatric donor cohort (aged 4-17 years) to capture the peak period of microbial diversity, with donor matching guided by a multidimensional algorithm and multi-omic quality control ^[21]^. Advanced cryogenic lyophilization and embedding technologies are employed to ensure microbial integrity and safety. Beyond this, we adopted a composite target of TIR >70% and TBR <4% for a more holistic assessment of efficacy and safety. However, our study is also limited by a relatively small sample size, single-center design, and relatively short(12-week) follow-up period, which might be insufficient to assess long-term glycemic benefit and sustained impact on residual islet β cell function.

Our findings will offer new clinical insight into the feasibility and effects of oral yFMT in adults with T1DM and provide the necessary evidence to power a subsequent multicenter large-scale RCT. Exploratory biomarker analyses conducted within this study may further pave the way for future individualized microbiome–based therapeutics.

## Supporting information

S1 File. SPIRIT 2025 Checklist.

S2 File. Protocol submitted for ethical approval. (Chinese)

S3 File. Protocol submitted for ethical approval. (English Translation)

Note:This document is an English translation of the protocol approved by the Ethics Committee. In case of discrepancies, the original Chinese version shall prevail.

## Data Availability

The data obtained during the course of this study will not be publicly available due to ethical and legal restrictions. De-identified data may be made available upon reasonable request to the corresponding author, subject to approval by the institutional ethics committee.

## Acknowledgments

We would like to express our sincere gratitude to all the participants and their families for their time and contribution to this study. Special thanks are extended to the healthy youth donors for providing the fecal microbiota samples. We are also grateful to the staff at the Department of Endocrinology and the Department of Clinical Nutrition at Zhongnan Hospital of Wuhan University for their support.

